# Longitudinal Sleep-Health Phenotypes Identified by Hierarchical Clustering in the Sleep Heart Health Study

**DOI:** 10.64898/2026.08.06.26359895

**Authors:** Antony Passaro, Kristy L. Meads, J. Kent Werner, Cameron H. Good

**Affiliations:** Thalamix Labs, LLC; Great Falls, Virginia, USA; Attune Neurosciences, Inc.; San Francisco, California, USA; Uniformed Services, University of the Health Sciences, MD, United States; Walter Reed National Military Medical Center, United States

**Keywords:** sleep health, hierarchical clustering, slow waves, aging, polysomnography, Sleep Heart Health Study, EEG, NREM sleep

## Abstract

Sleep health reflects interacting demographic, clinical, micro- and macroarchitectural, and neurophysiological factors that may not be captured by single metrics or diagnostic categories. We applied hierarchical clustering to longitudinal Sleep Heart Health Study data from 1,468 adults with complete polysomnographic, demographic/clinical, and pre-sleep electroencephalographic data at two visits separated by 5.19 ± 0.27 years. Forty nonredundant features selected from candidate demographic/clinical, sleep-stage, and pre-sleep spectral measures were clustered independently at each visit. Four reproducible sleep-health phenotypes emerged: a group with preserved deep sleep and favorable mental-health ratings; a large light-sleep group with low N3 and high N1; an older, physically unhealthy group with shorter total and rapid-eye-movement sleep; and a younger, physically healthy group with longer total and rapid-eye-movement sleep. The same population-level structure was evident at both visits, although only 37.7% of participants retained the same cluster assignment, with transitions most directed toward the light-sleep phenotype. An independent analysis of slow-wave morphology, excluded from cluster construction, differentiated all four phenotypes after false-discovery-rate correction. Groups with preserved or healthier sleep showed more numerous, higher-amplitude, steeper, and shorter slow waves, whereas the light-sleep and physically unhealthy groups showed weaker and more prolonged slow waves. Pre-sleep spectral features did not differ significantly across clusters after correction. These findings identify reproducible but individually dynamic sleep-health phenotypes and demonstrate that macro-architectural cluster structure is reflected in independent measures of NREM sleep microarchitecture.

**STATEMENT OF SIGNIFICANCE:** Sleep health is multidimensional, yet most studies evaluate isolated sleep variables or categorical disorders. We identified four reproducible sleep-health phenotypes across two examinations of the same adults separated by approximately five years. Although the population-level phenotype structure was stable, most individuals changed groups over time, commonly toward a light-sleep profile as individuals aged. Independent slow-wave measures strongly differentiated the phenotypes, whereas pre-sleep waking spectral measures did not. These findings show that sleep-health profiles are biologically meaningful but individually dynamic. Future work should determine which modifiable exposures drive transitions and whether phenotype-guided interventions can preserve deep sleep and broader health during aging.

## INTRODUCTION

Sleep-related changes across the lifespan are paralleled by alterations in key health metrics, reinforcing the relationship between sleep and systemic physiology. Poor sleep quality in midlife can predict the onset of chronic health conditions later in life, including hypertension, diabetes, and dementia.^1,2^ Insufficient or poor-quality sleep at any stage of life has been linked to a variety of negative health outcomes, including increased risk of metabolic disorders, cardiovascular disease, cognitive decline, and mental health issues.^3–6^ Moreover, changes in sleep architecture, such as a decline in deep sleep and rapid eye movement (REM) sleep, have been associated with an increased risk of mood disorders, accelerated aging process, cognitive decline, and immune function,^7–9^ while insufficient deep sleep has been linked to obesity and cardiovascular disease.^10^ Declines in sleep continuity and slow-wave activity are consistently linked with rising blood pressure, impaired glucose tolerance, and increased body mass index, all of which highlight sleep’s critical role in metabolic regulation.^11–13^ Similarly, reductions in REM sleep and increases in sleep fragmentation have been associated with heightened inflammatory markers and reduced autonomic flexibility, both of which predict greater cardiovascular morbidity with age.^14,15^ These findings underscore that sleep is not only shaped by aging processes but also contributes to the progression of chronic disease, with subtle alterations in sleep architecture and quality producing measurable effects on long-term health trajectories. Together, these findings highlight the necessity of integrating health indicators with sleep metrics to capture the multidimensional consequences of sleep aging.

By identifying and categorizing distinct sleep patterns and features, or clusters, we can characterize individual trajectories of sleep and health. Clustering serves as a powerful method for extracting meaningful patterns from large datasets, particularly by uncovering natural groupings based on shared characteristics without requiring predefined labels. Among clustering methods, hierarchical clustering is especially advantageous when data relationships are complex or layered. It constructs a nested tree-like structure, or dendrogram, that represents data similarities across multiple levels, enabling researchers to examine both broad categories and finer subgroups within datasets.^16^ The utility of cluster analysis in sleep research lies in its ability to illuminate complex relationships between physiological signals, behaviors, health, and distinct sleep profiles that may not be apparent when examining isolated metrics. For example, clusters of sleep profiles correlated with specific health risks, emphasizing the importance of tailored interventions.^17^ Similarly, it has been demonstrated that distinct sleep patterns could predict metabolic health, suggesting that cluster-based approaches could significantly enhance our understanding of sleep health.^10^ Moreover, these insights can inform personalized interventions that help transition individuals toward healthier sleep patterns before significant health deterioration occurs.^18,19^

In this study, we conducted a comprehensive cluster analysis of demographic and clinical measures, sleep macroarchitecture, and pre-sleep waking electroencephalography (EEG) features in two longitudinal assessments of the Sleep Heart Health Study (SHHS) cohort. The SHHS is a large, community-based, multicenter study designed to characterize sleep-disordered breathing and its cardiovascular and other health consequences. We tested whether reproducible multidomain sleep-health phenotypes emerged at both visits, quantified individual transitions between phenotypes, and independently evaluated the resulting clusters using slow-wave morphology that was not included in cluster construction.

## METHODS

### Participant database

The open-source SHHS database includes an initial examination (SHHS-1) of 5,793 participants who underwent unattended home polysomnography (PSG) between November 1995 and January 1998 and a follow-up examination (SHHS-2) of 2,651 returning participants between January 2001 and June 2003 (mean interval, 5.19 ± 0.27 years).^20,21^ The SHHS was a multicenter cohort study sponsored by the National Heart, Lung, and Blood Institute to examine associations between sleep-disordered breathing and cardiovascular and other health outcomes. For the present analysis, only participants with data from both examinations were considered. Data collection was conducted in participants’ homes using standardized unattended overnight polysomnography, with equipment setup and calibration performed by trained technicians. The Compumedics P-Series portable system recorded two EEG derivations (C3-A1 and C4-A1), electrooculography, electromyography, electrocardiography, airflow, respiratory effort, oximetry, body position, and ambient light. Studies were transferred to a central Reading Center for manual scoring and quality review. Demographic, clinical, questionnaire, and laboratory measures were also obtained.^20,22^

Among the 2,651 returning participants, 918 were excluded because one or more required demographic/clinical, EEG, or polysomnographic values were missing, and 223 were excluded because at least one required metric had a zero value that was incompatible with the planned ratio-based analyses. This yielded 1,588 participants with complete candidate data. Each of the 40 retained metrics was then normalized separately within SHHS-1 and SHHS-2, and 120 participants with at least one absolute z-score greater than 7 were excluded. The final sample included 1,468 participants with an estimated mean age of 62.5 ± 11.9 years at SHHS-1 and 68.0 ± 12.2 years at SHHS-2.

### Sleep and health metrics for cluster analysis

Feature selection was performed using SHHS-1. Candidate variables comprised of 250 pre-sleep EEG features, 937 demographic and behavioral measures, and approximately 1,000 polysomnographic features, including sleep-stage counts, percentages, and stage-ratio measures. Principal component analysis was first applied separately within each feature domain, retaining components that together explained more than 90% of variance. Features with the largest contributions to the retained components were then selected, yielding 65 candidate features. To reduce redundancy, features that were highly intercorrelated with another feature in the same domain were removed, resulting in 40 variables. This fixed 40-feature set was subsequently used for hierarchical clustering in both SHHS-1 and SHHS-2.

The top 40 identified metrics from the SHHS-1 dataset are listed in **Table S1**, and include 15 demographic, clinical, and laboratory (DCL), 12 PSG and 13 EEG. The 15 DCL metrics include the Respiratory Disturbance Index (*RDI*), which is a measure of obstructive sleep apnea (OSA) severity and reflects the average number of apneas plus hypopneas observed per hour of sleep. *SystBP* and *DiasBP* are systolic and diastolic blood pressure, respectively. *NECK* is a measure of neck circumference in centimeters. *Sleepy* refers to a specific question asking how often an individual feels sleepy during the day with a response value of 1 to 5 [1: Never, 2: Rarely (1x/month or less), 3: Sometimes (2-4x/month), 4: Often (5-15x/month), 5: Almost Always (16-30x/month)]. This question for the SHHS dataset has been shown to relate to nocturia (waking up from sleep for urination) and OSA.^23,24^ The subjective body pain (*bp*) index is taken from the medical outcomes study short form 36 (SF-36) to assess overall body pain [10 to 100] with a higher value corresponding to “no pain or limitations due to pain in the past 4 weeks”.^25,26^ The general health (*gh*) perception scale is based on 5 questions around perceived general health as part of the SF-36 [10 to 100] with a higher score corresponding to the belief that the subject is in excellent health. The general mental health (*mh*) index is from the SF-36 [10 to 100] based on 5 questions with a higher score corresponding to feeling “peaceful, happy, and calm all of the time in the past 4 weeks”. The *mcs* and *pcs* metrics refer to the mental and physical summary component scales from the SF-36 [10 to 100] with higher values corresponding to better health in the respective areas. *Gender, age, body mass index (BMI), weight, and height* refer to their respective demographics and body measurements.

Twelve PSG-derived metrics were computed for both nights of sleep for each subject. *Total sleep time* (TST) is a count of the number of 30s, non-overlapping, epochs that were labeled as one of the 4 stages of sleep (*N1, N2, N3, REM*). The same approach applies to the following metrics as well: *N1 count, N3 count, REM count (note that N2 count was not a top feature in the PCA down-selection)*. *Sleep percentage* is the percent of the total nightly recording that was labeled as one of the four sleep stages (i.e. not waking). Both *REM percentage* and *N3 percentage* are the respective percentages relative to all epochs labeled as a sleep stage. The following metrics are ratios of sleep stages based on epoch count: *N2/N3, N1/REM, N3/REM*. Two complex sleep stage ratios were also utilized: (*N2+N3)/REM* was calculated by summing the count of all N2- and N3-labeled epochs and dividing by the count of REM-labeled epochs and *REM/N3/N1* was calculated as the number of REM-labeled epochs divided by the number of N3-labeled epochs, which is further divided by the number of N1-labeled epochs.

Thirteen pre-sleep EEG metrics were computed from the C3-A1 derivation using only epochs scored as wake before sleep onset. Sleep onset was operationally defined as the first 30-second epoch scored as any sleep stage (N1, N2, N3, or REM). Restricting EEG features to the preceding waking period reduced overlap with the sleep-stage variables used elsewhere in the model. A minimum of 10 minutes of pre-sleep wake data was required; among included participants, the available pre-sleep wake interval averaged 62.1 ± 40.42 minutes. For each nonoverlapping 30-second wake epoch, a fast Fourier transform was used to estimate spectral power, which was normalized by mean power from 0.1 to 30 Hz. Band ratios were included because relative spectral relationships can capture arousal-related and state-related EEG variation and have also been used in automated sleep-stage classification (Stephansen et al., 2018). Spectral bands were defined as follows: d1, 0.1-0.5 Hz; d2, 0.5-1.5 Hz; t1, 4-8 Hz; a1, 8-10 Hz; a2, 10-13 Hz; a3, 8-13 Hz; b1, 13-20 Hz; and b2, 20-30 Hz. Five ratios were calculated: d2/d1, b1/b2, d3/d2, t1/d1, and beta-plus-theta/all-alpha (bta).

### Hierarchical clustering

Hierarchical clustering was employed to identify patterns within the 40-dimension sleep dataset using Ward’s linkage method. To begin, the Euclidean distance metric was calculated for all pairs of data points to quantify dissimilarities between observations. Subsequently, Ward’s linkage was utilized as the agglomerative criterion, focusing on minimizing the total within-cluster variance at each step of the clustering process by calculating the within-cluster sum of squares based on Euclidean distance. This method iteratively merged pairs of clusters, selecting the pair that resulted in the smallest increase in the total within-cluster variance. The process continued until all data points were merged into a single cluster, producing a hierarchical tree structure (dendrogram) that represented the nested grouping of observations, allowing for the identification of natural groupings within the multidimensional sleep dataset. Branches on the dendrogram indicate the points at which clusters were merged during the agglomerative process, with their lengths reflecting the Euclidean distances between merged clusters and highlight their level of similarity. Each leaf at the bottom of the dendrogram corresponds to an individual data point.

### Cluster evaluation

The silhouette criterion was applied using correlation as the distance metric to determine the optimal number of clusters found in the datasets. The silhouette score measures how similar an object is to its own cluster compared to other clusters, providing an assessment of clustering quality. For each data point, the silhouette coefficient was calculated as the difference between the average distance to points within the same cluster (cohesion) and the average distance to points in the nearest neighboring cluster (separation), normalized by the maximum of these two values. Correlation was used as the distance metric to account for the relationships between variables in the 40-dimensional sleep dataset. The number of clusters that maximized the average silhouette score was selected as the optimal clustering solution. This approach was independently applied to both datasets (SHHS-1 and SHHS-2), as well as the two datasets appended together (Figure S1).

### Dimensionality reduction and visualization

Principal component analysis was applied to the 40-dimensional sleep dataset to visualize the clustering results in a reduced dimensional space. The first three principal components, which together explained most of the variance in the dataset, were extracted to create a three-dimensional component space. The data points were then plotted in this reduced space, with each point colored according to its cluster assignment (Figure 1).

**Figure 1.**
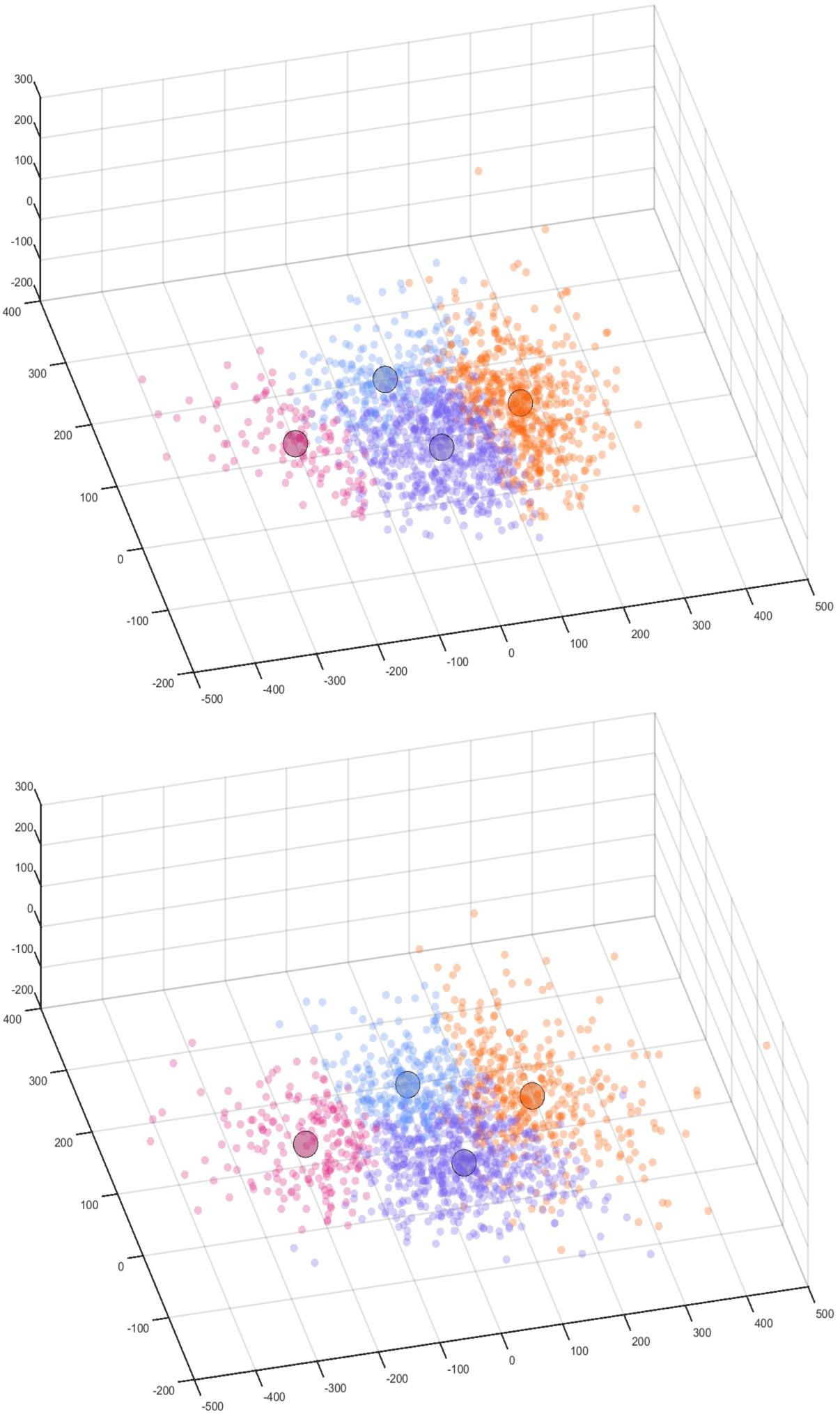
Principal component analysis scatter plots of clustering results for SHHS-1 and SHHS-2 in a reduced 3-dimensional component space. The color-coded clusters represent the four identified groups: Cluster 1 (blue) – mentally healthy with good deep sleep; Cluster 2 (purple) – poor deep sleep; Cluster 3 (pink) – physically unhealthy and older group with poor overall sleep; and Cluster 4 (orange) – physically healthy, younger group with good sleep. The plots demonstrate clear separation between the clusters along two axes, with Clusters 1 and 2 extending in opposite directions and Clusters 3 and 4 following a similar pattern. The similarity of the clustering patterns across both datasets highlights the robustness of these distinct groups across the SHHS cohort.

### Statistical methods

We conducted a series of one-way analyses of variance (ANOVAs) with cluster assignment as the independent factor to evaluate differences across clusters for DCL, sleep, and EEG features. Each feature was tested independently, yielding a p-value for the null hypothesis that the mean value did not differ across the four clusters. Given the large number of comparisons, we implemented the False Discovery Rate (FDR) to correct for multiple comparisons and control for Type I errors,^27^ with statistical significance defined as q < 0.05. One-way ANOVAs with FDR correction were similarly applied to each slow-wave metric to evaluate for differences across clusters within each of the two SHHS datasets.

The results of the cluster analyses were visualized using violin plots, providing a combined view of the distribution, density, and variability of each metric across clusters. Cluster-level means ± standard error of the mean (SEM) were overlaid as black points to highlight central tendencies against the backdrop of the full distribution. This approach allowed us to simultaneously assess statistical significance, effect size, and distributional characteristics of each feature across clusters. Features meeting the FDR threshold are marked with asterisks in the plots. By integrating violin plots with FDR-corrected ANOVA testing, we captured both robust statistical differences and distributional nuances between clusters, thereby strengthening the interpretability of multidomain comparisons across DCL, sleep, and EEG metrics.

### EEG artifact rejection

Artifact rejection is a critical component of ensuring data quality. We developed and implemented a custom automated artifact detection procedure in which the power spectra from each epoch were log-transformed using the base-10 logarithm. An artifact threshold was then computed using the formula where SP denotes the spectral power in each respective frequency band:

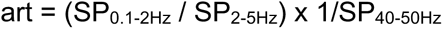

Epochs exceeding a threshold value of 7 were classified as artifactual and excluded from further analysis. This method effectively removed segments contaminated by low-frequency drift or high-frequency noise (e.g., muscle activity), enhancing the reliability of spectral estimates used in downstream analyses.

### Slow wave analysis

Slow waves were identified independently within N2 and N3 sleep and were not used to construct the clusters. Candidate events began at a negative-going zero crossing and included a contiguous negative half-wave lasting 500-4,000 ms with a negative peak of at least -20 µV. The subsequent positive half-wave was included to calculate peak-to-peak amplitude. Events were retained when total peak-to-peak amplitude exceeded 40 µV. The lower amplitude threshold, relative to the 75- µV criterion commonly used for classic high-amplitude slow waves, was selected to retain lower-amplitude but physiologically relevant events expected in an older community cohort.^28,29^ The 4,000-ms upper bound excluded prolonged nonoscillatory deflections.

For each detected slow wave, we quantified eight complementary metrics to capture different aspects of slow wave morphology and dynamics: (1) Count, the total number of slow waves; (2) Density, the number of slow waves per minute of N2/N3 sleep; (3) Negative Amplitude, the minimum negative peak value (µV); (4) Total Amplitude, the peak-to-peak amplitude (µV); (5) Duration, the total time from negative onset to positive offset (ms); (6) Negative Duration, the length of the negative half-wave (ms); (7) DownSlope, the steepest negative-going slope (µV/s); and (8) UpSlope, the steepest positive-going slope (µV/s). Together, these measures provide a comprehensive description of both the abundance and morphology of slow waves across clusters.

## RESULTS

### Cluster identification and structure

Hierarchical clustering combined with the silhouette criterion was applied independently to both the SHHS-1 and SHHS-2 datasets, and a combined dataset (SHHS-1 + SHHS-2). Results identified four clusters as the optimal solution for SHHS-1 and the combined datasets, while two primary clusters were found to be optimal for SHHS-2 that further divided into the same four-cluster solution (Figure S1). Tables S1 and S2 show the number of participants associated with each cluster for SHHS-1 and SHHS-2, respectively. Additionally, the tables present the summarized values for each feature by cluster, with statistically significant differences across clusters indicated in bold following ANOVA with FDR correction. Notably, the four identified clusters were consistent across both datasets, underscoring the stability of this sleep phenotype classification approach.

We assigned descriptors to each cluster according to their most distinguishing demographic, health, and sleep-stage features. Cluster 1 (mentally healthy) showed the strongest deep sleep profile, with the highest N3 count and percentage, as well as the highest mental health scores. Cluster 2 (light sleep), the largest group, had average demographic characteristics but was distinguished by the lowest N3 percentage and the highest N1 count, reflecting a predominance of lighter sleep stages. The prominence of this group highlights that light, non-restorative sleep may be especially common in older adults, consistent with prior reports linking diminished N3 sleep to widespread superficial sleep patterns and poorer health outcomes.^8,30^ Cluster 3 (physically unhealthy) was the oldest group and displayed the poorest overall health, with the lowest general health ratings, heaviest body weight, and the least total and REM sleep. In contrast, Cluster 4 (physically healthy) was the youngest group, characterized by the greatest total sleep time and REM sleep, along with more favorable physical health measures.

Three-dimensional PCA revealed distinct separation between the four clusters in each dataset, as each cluster extended in opposite directions along two principal axes to create opposing clusters (Figure 1). Specifically, Cluster 1 (blue) was positioned opposite Cluster 2 (purple), reflecting the differences in deep sleep quality between these groups. Cluster 3 (pink), the older group characterized by poor overall sleep and worse physical health, was located opposite Cluster 4 (orange), the younger group with better physical health and better sleep metrics. This opposition of clusters suggests that distinct sleep and health profiles map onto clear and separable dimensions, much like the opposing phenotypes identified in prior sleep studies using clustering methods.^31,32^ Further, these PCA findings were consistent across SHHS-1 and SHHS-2 (Figure 1), indicating that the opposing relationships between these clusters were stable over time (years).

Dendrogram analyses further supported the distinct hierarchical structure of the clusters, with similar branching patterns observed across SHHS-1 and SHHS-2 (Figure 2). In SHHS-1, Cluster 3 (pink), which represented the physically unhealthy group with poor overall sleep, formed its own distinct branch, emphasizing its unique position as the group with the worst health and sleep outcomes. This result suggests that poor health, particularly in older individuals, is closely tied to poor sleep quality, in line with previous research showing strong correlations between aging, poor physical health, and reduced sleep efficiency.^1,33^

**Figure 2.**
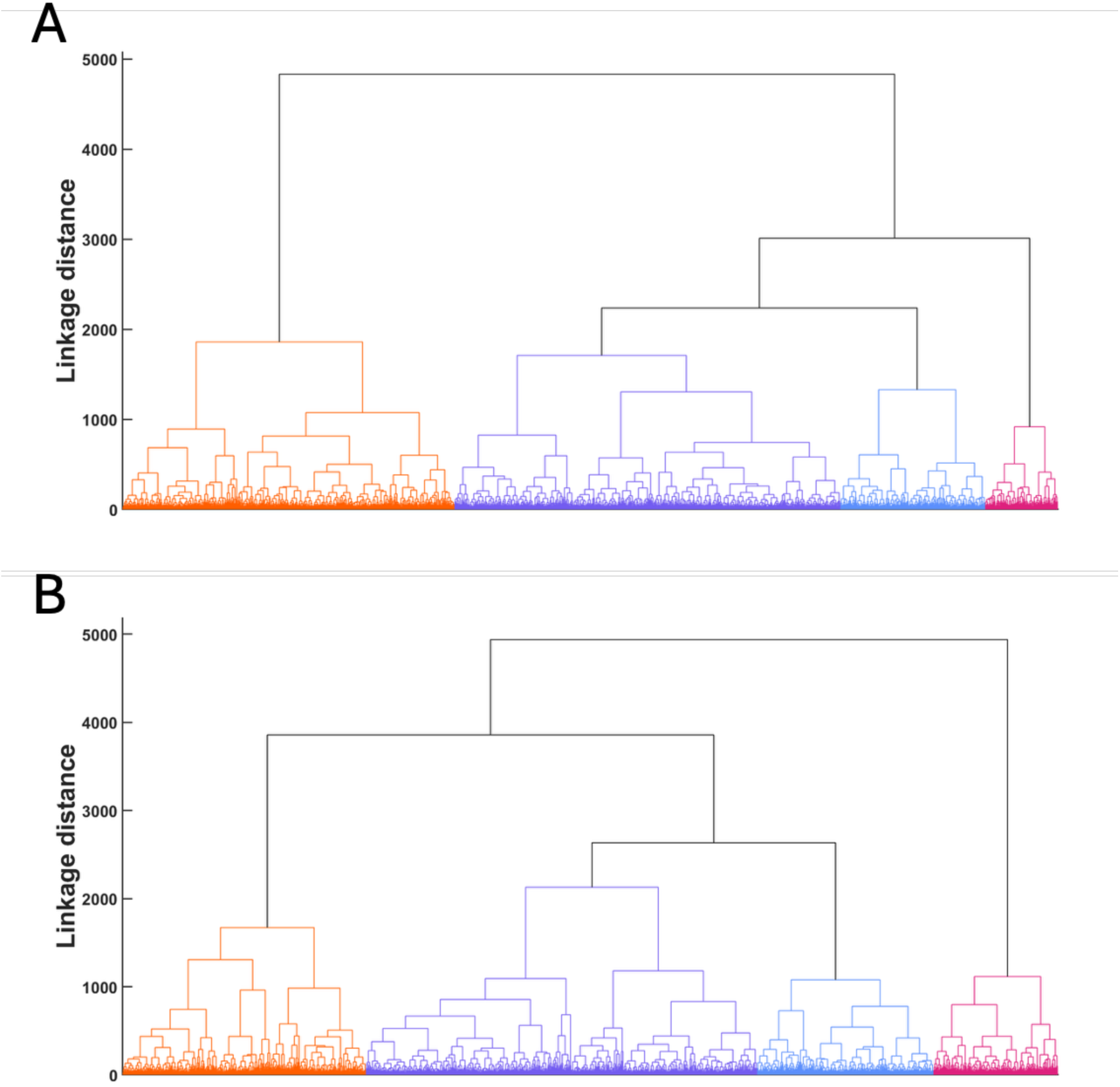
Dendrograms of hierarchical clustering for SHHS-1 (A) and SHHS-2 (B). Dendrograms displaying the hierarchical clustering structure using Ward’s linkage method. Both dendrograms reflect the four-cluster solution identified through the silhouette criterion and PCA analysis. Each branch represents the merging of data points or clusters, with branch lengths corresponding to the degree of dissimilarity (Euclidean distance) between clusters. The clusters were color-coded based on their final group assignments, providing a visual summary of the hierarchical clustering structure and the relative distances between clusters. **(A)** In SHHS-1, Cluster 3 (pink) – the physically unhealthy, older group – forms a distinct group early in the clustering process, separating from the other clusters. Clusters 1 (blue) and 2 (purple) – mentally healthy with good deep sleep and poor deep sleep, respectively – remain closely related, while Cluster 4 (orange) – physically healthy with good sleep – merges later. **(B)** In SHHS-2, Cluster 4 (orange) – the physically healthy, younger group with good sleep – forms its own distinct branch early, separating from the other three clusters. The structure is slightly different compared to SHHS-1, with the poor sleep Cluster 2 (purple) showing tighter connections with the remaining groups.

In SHHS-2, the hierarchical structure showed that Cluster 4 (orange), the group characterized by younger age and good sleep, became its own separate group, distinct from the other three clusters. This suggests that, over time, this cluster retained its identity more clearly than others, potentially due to the stability of good sleep in younger, healthier individuals. Interestingly, this mirrors findings in longitudinal studies where good sleep patterns in early adulthood often predict better health outcomes in later life.^34,35^ In contrast, the other clusters, particularly those with poorer sleep, may be more prone to transitions as health and sleep patterns naturally deteriorate with age.

### Cluster feature weights

The correlation between each feature and the cluster groupings was calculated to identify the features most strongly associated with each cluster. Specifically, the relationship between each of the 40 features and the assigned clusters was quantified using correlation coefficients, which measured the strength and direction of association. Features with the highest weights and statistically significant adjusted p-values (FDR < 0.05) were reported as the top distinguishing characteristics of each cluster (Figure 3).

**Figure 3.**
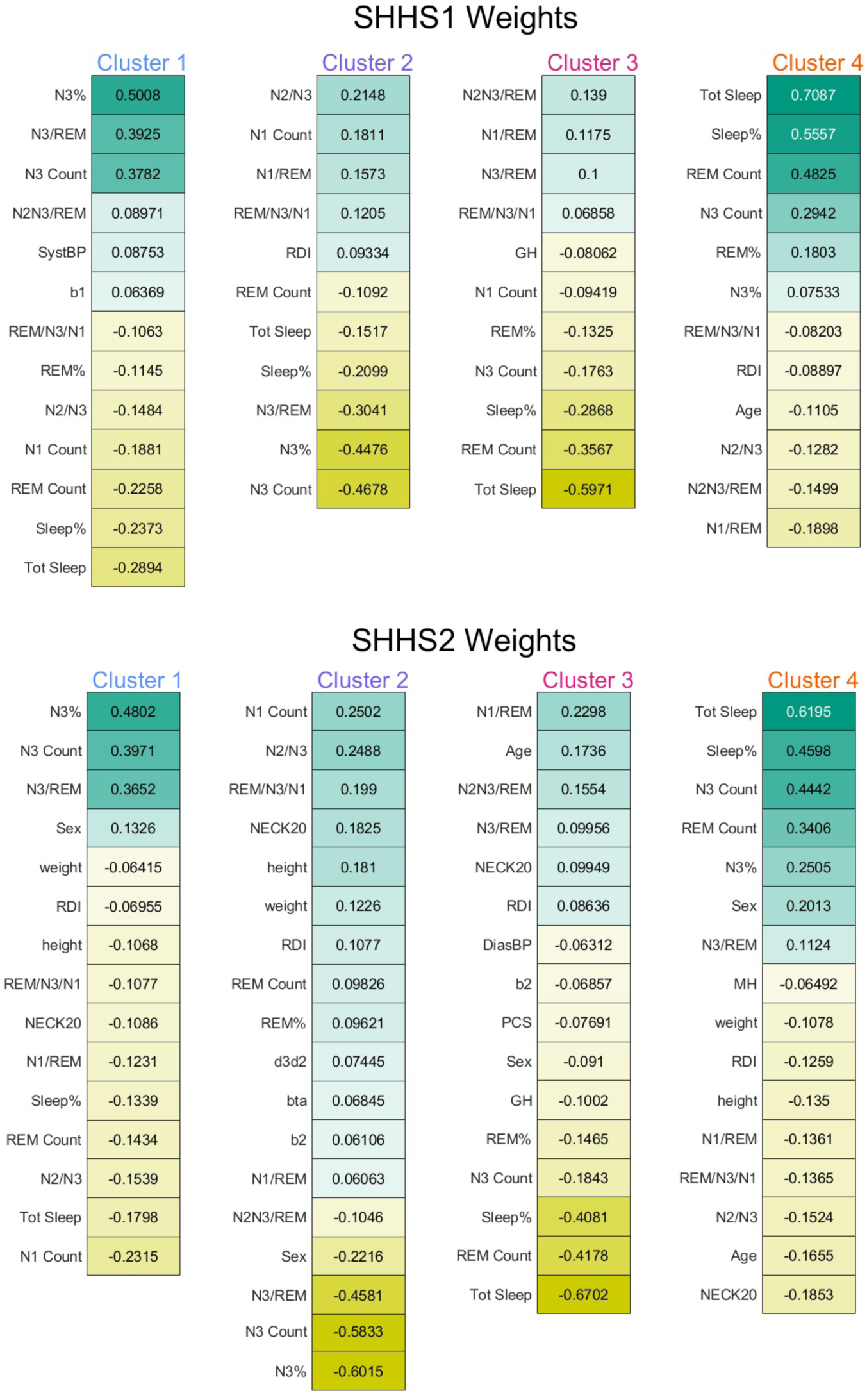
Statistically significant feature weights for SHHS-1 (top) and SHHS-2 (bottom) after false-discovery-rate correction. Columns display features significantly associated with each cluster, ordered by weight. Positive and negative weights indicate the direction of association between a feature and cluster membership; shading intensity indicates weight magnitude. N3-related features most strongly distinguished Clusters 1 and 2, whereas total sleep time, sleep percentage, and REM-related measures primarily distinguished Clusters 3 and 4. The broad pattern was reproducible across visits, with modest differences in feature ordering and magnitude.

Feature weight analysis revealed that Clusters 1 (mentally healthy) and 2 (light sleep) were primarily differentiated by opposing N3-related features. Cluster 1 was positively defined by higher N3 percentage, N3 count, and N3/REM ratio, underscoring the central role of deep sleep as a marker of healthier sleep. In contrast, Cluster 2 was negatively weighted on these same features, with the strongest contributions coming from lower N3 values and higher N1 counts. This pattern highlights how reduced N3 and elevated N1 characterize lighter, less restorative sleep, consistent with prior work linking diminished deep sleep to adverse health and aging outcomes.^8,30,36^

By comparison, Clusters 3 (physically unhealthy) and 4 (physically healthy) were distinguished by opposing features related to overall sleep quantity and REM sleep. Cluster 3 was most strongly negatively weighted on total sleep time and REM count, reflecting insufficient sleep and poorer general health, while Cluster 4 showed the opposite pattern, with high positive weights for total sleep, sleep percentage, and REM count, consistent with a more robust and healthier sleep profile. In SHHS-2, Cluster 4 also showed N3 count as a strong positive contributor, suggesting that deep sleep complemented REM in defining this healthier group. These opposing weight patterns indicate that while N3 sleep is the critical differentiator for some clusters, overall sleep duration and REM are more salient for others, aligning with prior research emphasizing the multifactorial nature of sleep health.^37^

### Demographic, Clinical, and Sleep Metric Differences

Z-scored DCL measures revealed significant differences across clusters, underscoring the multidimensional nature of these groups (Figure S2A). Several variables reached statistical significance after FDR correction, including age, sex distribution, RDI, and blood pressure. Cluster 3 (pink) was consistently the oldest and least healthy group, whereas Cluster 4 (orange) was the youngest and most physically healthy. Neck circumference, weight, and self-reported general health also differed significantly between clusters, with Cluster 2 (purple) showing higher neck circumference and poorer health ratings compared to Cluster 1 (blue) and Cluster 4. These patterns are consistent with prior reports linking older age, higher RDI, and adverse cardiometabolic factors to impaired sleep and health outcomes.^38,39^

Sleep metrics also demonstrated robust and statistically significant differences between clusters (Figure S2B). Cluster 1 (blue), characterized by strong restorative sleep, showed significantly higher N3 count and REM count, indicating preserved deep and REM sleep. In contrast, Cluster 2 (purple), the poor deep sleep group, had the highest N1 count and the lowest N3 and REM counts, consistent with light, fragmented sleep and reduced restorative quality. Cluster 3 (pink) also showed significantly lower N3 and REM counts than Clusters 1 and 4, reinforcing its profile as an older, less healthy group with disrupted sleep architecture. By comparison, Cluster 4 (orange) demonstrated intermediate to high values for N3 and REM sleep and relatively low N1 counts, consistent with resilient, healthy sleep. These findings parallel prior literature linking reduced N3 and REM sleep with impaired cognitive and physiological outcomes, and elevated N1 with poor sleep consolidation.^40,41^

EEG spectral metrics from the pre-sleep awake period showed no statistically significant differences after FDR correction (Figure S2C). However, qualitative trends were observed: Cluster 3 (pink) tended to exhibit elevated delta and theta activity, while Clusters 1 (blue) and 4 (orange) showed relatively stronger alpha–beta power compared to Clusters 2 and 3. While these differences did not meet statistical significance, they are directionally consistent with prior studies linking increased waking low-frequency power to aging and reduced cortical integrity, and stronger alpha–beta rhythms to healthier neural function.^42,43^

### Sleep features across clusters

Key sleep metrics visually reinforced the distinctions between the four clusters in both datasets. Radar plots (Figure 4) show that Cluster 1 (mentally healthy, blue) and Cluster 2 (light sleep, purple) were nearly opposite to one another, with Cluster 1 defined by higher N3 measures and Cluster 2 by lower N3 and higher N1, underscoring their contrasting deep sleep characteristics. Similarly, Cluster 3 (physically unhealthy, pink) and Cluster 4 (physically healthy, orange) displayed opposite profiles, with Cluster 3 characterized by lower total sleep and REM measures, and Cluster 4 by higher values on these metrics, consistent with poorer versus healthier overall sleep.

**Figure 4.**
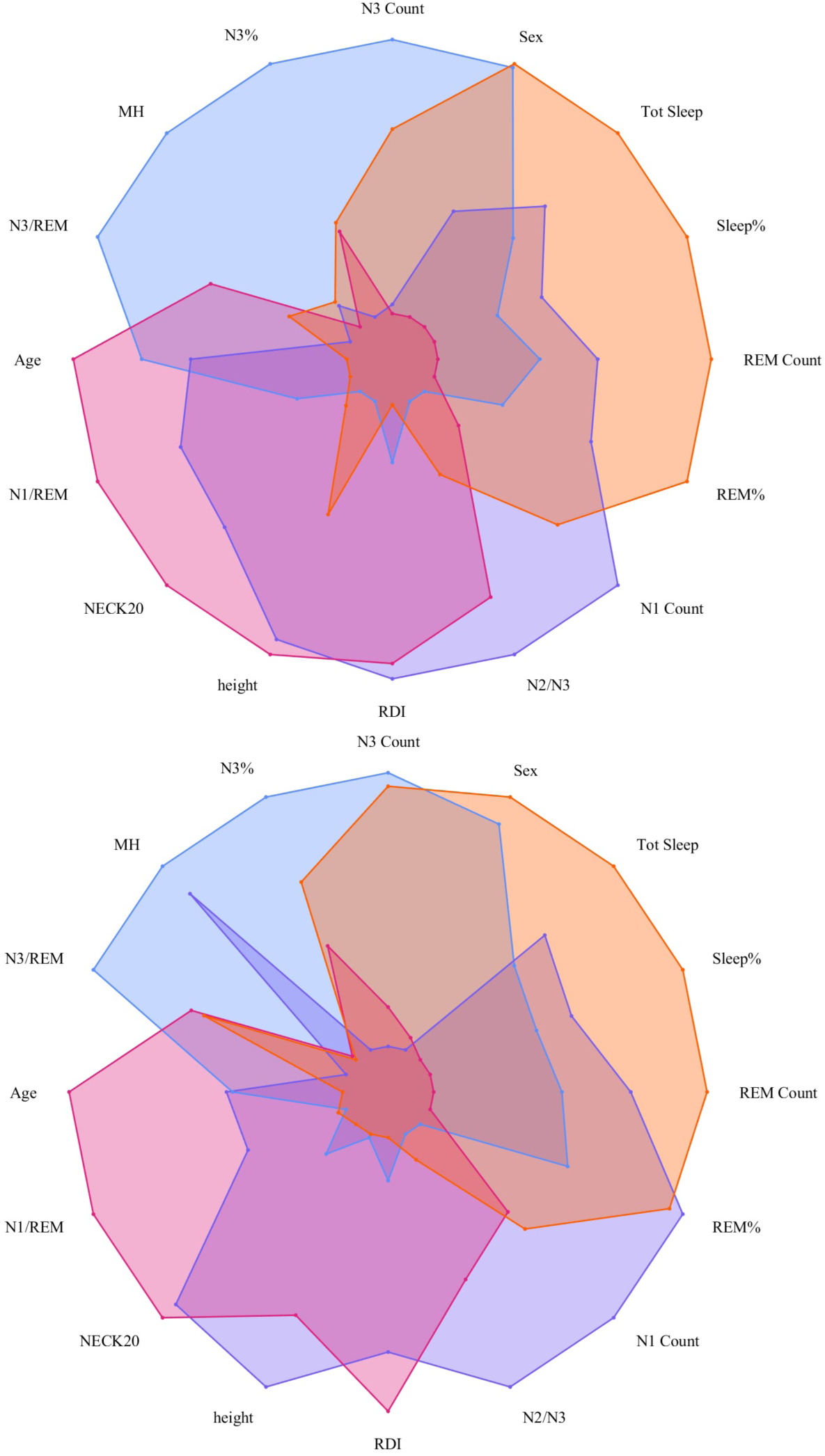
Radar plots of top cluster weights for key metrics. Plots illustrate the top distinguishing features for each cluster based on feature weights. Each axis corresponds to a sleep or health metric, with values normalized for comparison. Cluster 1 (Mentally Healthy, blue) and Cluster 2 (Light Sleep, purple) appear opposite to one another, reflecting their contrasting N3- and N1-related sleep profiles. Cluster 3 (Physically Unhealthy, pink) and Cluster 4 (Physically Healthy, orange) also show opposing patterns, with Cluster 3 negatively weighted on total sleep and REM measures and Cluster 4 positively weighted on these same features.

Despite these oppositions, areas of overlap were also evident. In both SHHS-1 (top panel) and SHHS-2 (bottom panel), Cluster 1 and Cluster 3 shared some similarities, particularly in N3-related features, while Cluster 2 and Cluster 4 showed partial overlap on sleep quantity measures. This suggests that the clusters, while distinct, are not entirely discrete categories; instead, they share intermediate profiles that reflect gradations in sleep and health. Such continuity aligns with prior work demonstrating that sleep phenotypes often exist along a spectrum, particularly in older populations.^44,45^ Together, these radar plots illustrate that sleep architecture, and health outcomes, are best understood as existing along a continuum rather than in binary states.

### Cluster transfer across SHHS datasets

To evaluate longitudinal stability of sleep–health profiles, participants were assigned unique index values and tracked across SHHS-1 and SHHS-2 to determine changes in cluster membership over time. Transitions between clusters were quantified by calculating the proportion of individuals who remained in the same cluster or moved to a different cluster at follow-up (mean interval 5.19 ± 0.27 years). These proportions were then used to generate a transfer diagram in which each cluster at baseline and follow-up was represented as a node, and connecting lines indicated the relative flow of participants between clusters (Figure 5). Line thickness and opacity were scaled to the percentage of participants undergoing each transition, and node size was scaled to the proportion of the cohort contained within each cluster. This approach allowed for a visual and quantitative representation of the stability and evolution of cluster membership across time, shedding light on the longitudinal evolution of sleep phenotypes.

**Figure 5.**
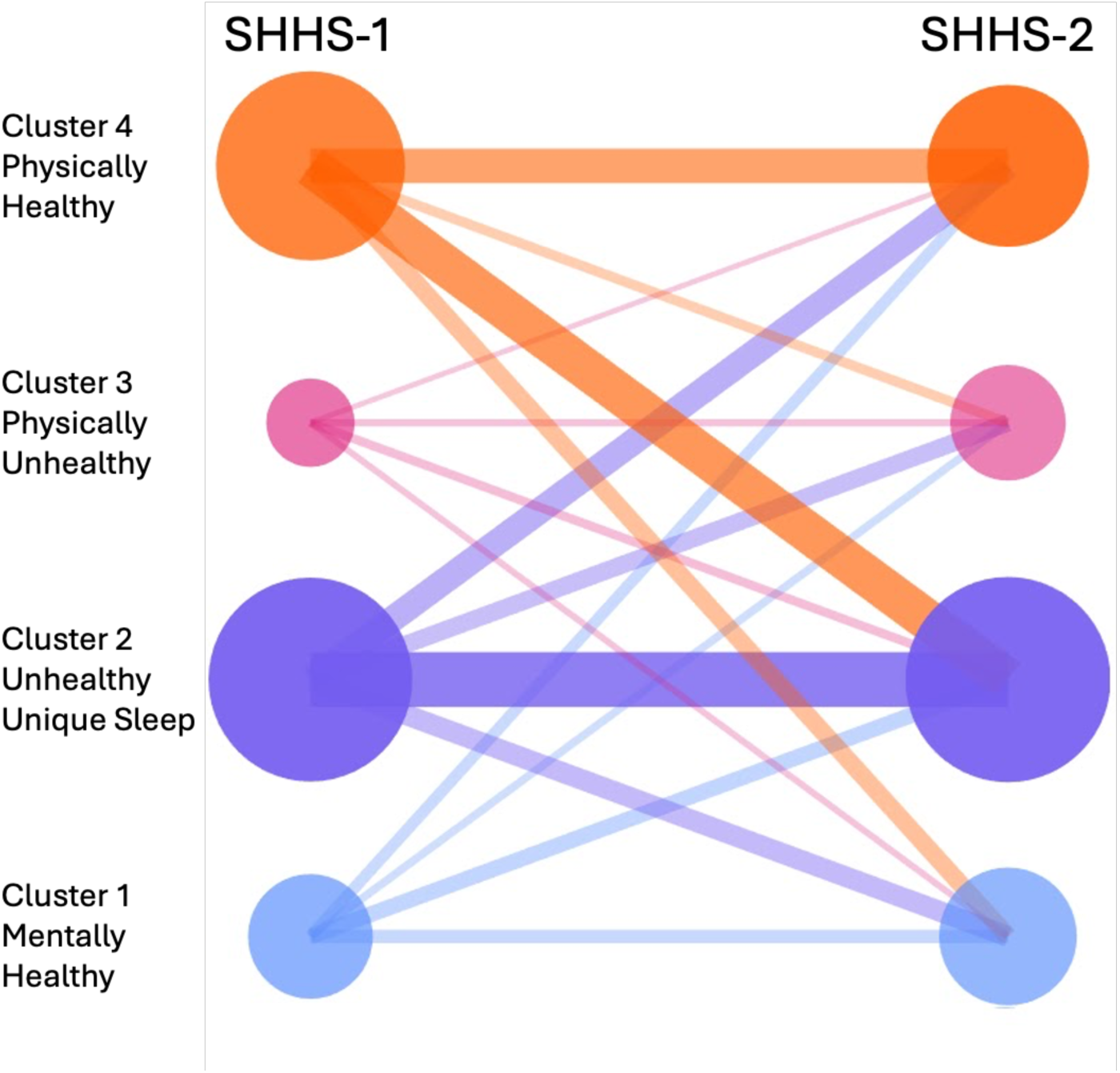
Changes in participant cluster membership from SHHS-1 to SHHS-2. Nodes represent the four clusters at each examination, and connecting lines represent participant transitions over the mean 5.2-year interval. Line thickness and opacity are proportional to the percentage of participants following each transition, and node size is proportional to cluster size. The corresponding counts and row percentages are provided in Table 1.

**Table 1.** Transition matrix of participant cluster assignments from SHHS-1 to SHHS-2. Each cell reflects the number of participants and the corresponding percentage (based on the SHHS-1 cluster row total) who transitioned to a given SHHS-2 cluster (columns). Diagonal cells represent stable cluster memberships across the two time points, while off-diagonal entries indicate transitions to different clusters over time. Of the 1468 total participants, Cluster 2 was the most stable (47.2% remained in the same cluster), while Cluster 1 showed more distributed transitions, with only 27.3% remaining in the same cluster and 72.7% transitioning to other groups.

| **SHHS-1 to SHHS-2** | **Cluster 1** | **Cluster 2** | **Cluster 3** | **Cluster 4** |
| --- | --- | --- | --- | --- |
| **Cluster 1** | 62 (27.3 %) | 76 (33.5 %) | 35 (15.4 %) | 54 (23.8 %) |
| **Cluster 2** | 102 (16.8 %) | 286 (47.2 %) | 86 (14.2 %) | 132 (21.8 %) |
| **Cluster 3** | 26 (22.8 %) | 39 (34.2 %) | 29 (25.4 %) | 20 (17.5 %) |
| **Cluster 4** | 87 (16.7 %) | 213 (40.9 %) | 45 (8.6 %) | 176 (33.8 %) |

Of the 1,468 participants, 553 (37.7%) remained in the same cluster from SHHS-1 to SHHS-2, indicating that individual sleep-health profiles were substantially more dynamic than the stable population-level four-cluster structure (Table 1). Cluster 2, characterized by low N3 and relatively preserved subjective health, showed the greatest stability: 286 of 606 participants (47.2%) retained this assignment. Cluster 1 showed lower stability, with 62 of 227 participants (27.3%) remaining in Cluster 1 and the largest subgroup, 76 participants (33.5%), transitioning to Cluster 2. This pattern is consistent with an age-related shift toward reduced deep-sleep expression, although the observational design does not identify the causes of individual transitions.

Cluster 4 (orange), considered the healthiest group based on physical metrics and subjective health ratings, retained 176 out of 521 participants (33.8 %), while a striking 213 individuals (40.9 %) transitioned to Cluster 2. This transition pattern suggests that even among the healthiest sleepers, vulnerability to deep sleep deterioration over time is common—a trend supported by aging literature showing marked N3 decline across adulthood.^8,30^ Interestingly, 87 individuals (16.7 %) from Cluster 4 transitioned to Cluster 1, and 45 (8.6 %) to Cluster 3, illustrating that although many remained resilient, a subset experienced broader declines in both sleep and physical health.

Cluster 3, the older group marked by poor overall sleep and health, was the smallest at SHHS-1 (n = 114). Twenty-nine participants (25.4%) remained in Cluster 3, 39 (34.2%) transitioned to Cluster 2, 26 (22.8%) transitioned to Cluster 1, and 20 (17.5%) transitioned to Cluster 4. Because cluster membership was recomputed independently at each visit, these transitions should be interpreted as changes in multivariate profile rather than direct evidence of clinical deterioration or improvement.

Overall, these patterns support a model of both resilience and decline in sleep-related health profiles. Transitions were not random but exhibited directional trends: individuals from healthier groups tended to move into poorer sleep quality profiles, especially Cluster 2, while those with mixed or poor profiles showed more variable outcomes. These findings are consistent with prior longitudinal research showing that deep slow wave sleep declines more predictably than other features of sleep architecture and that recovery from entrenched poor sleep is relatively rare.^8,46^ The identification of these transition trajectories has important implications for targeted intervention—particularly in preventing decline among otherwise healthy sleepers and understanding which factors may contribute to rare cases of improvement.

### Independent slow wave analysis

Analysis of slow wave features, which were not part of the original clustering procedure, revealed consistent statistical differences across the four identified clusters, further highlighting distinct physiological sleep profiles that extend into micro-architectures (Figure 6). ANOVA with FDR correction revealed robust differences for each metric in both SHHS-1 and SHHS-2 (Table S3). Cluster 1 (blue), previously characterized by robust deep sleep, exhibited the highest total amplitude slow wave values among all groups, reflecting strong and consolidated oscillations. This group also showed relatively steep down- and up-slopes, as well as the shortest slow wave durations, consistent with efficient and synchronous oscillatory dynamics. Such features align with prior studies showing that greater slow wave amplitude and steeper slopes are markers of healthy, resilient sleep physiology.^47,48^

**Figure 6.**
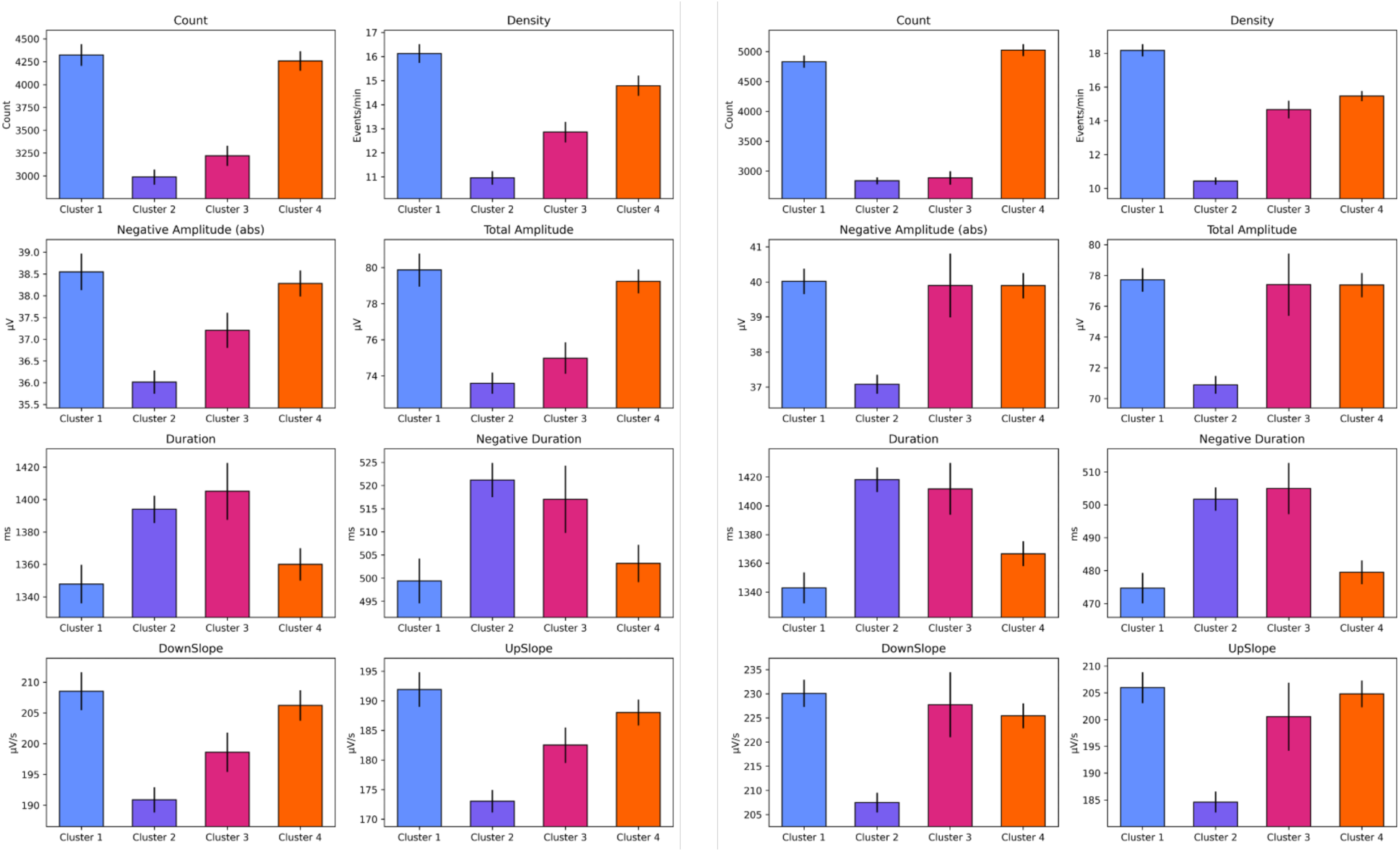
Slow-wave metrics across clusters in SHHS-1 (left) and SHHS-2 (right). Distributions of slow-wave count, density, negative amplitude, total amplitude, duration, negative duration, downslope, and upslope for each cluster. Bar graphs and error bars indicate cluster means ± SEM. All metrics statistically differed across clusters after false-discovery-rate correction in both datasets. Clusters 1 and 4 generally showed more abundant, higher-amplitude, steeper, and shorter slow waves, whereas Clusters 2 and 3 showed weaker and more prolonged slow-wave morphology.

In contrast, Cluster 2 (purple), representing light and fragmented sleep, showed the lowest slow wave counts, density, down- and up-slopes, and amplitudes, combined with the longest slow wave durations. This profile reflects weak and prolonged oscillations, indicative of disrupted synchrony in cortical networks during non-REM (NREM) sleep. These characteristics are consistent with findings linking reduced slow wave density and flatter slopes to aging and poorer sleep quality.^48,49^

Cluster 3 (pink), associated with older and physically less healthy individuals, also showed diminished slow wave activity. Specifically, this cluster displayed very low slow wave counts just above Cluster 2, the second lowest total amplitudes, and long durations, further suggesting inefficient or dysregulated slow oscillations. Prior studies have reported similar associations between aging, health decline, and reductions in slow wave amplitude and temporal precision.^8,42^

Finally, Cluster 4 (orange), corresponding to the younger and physically healthy group, demonstrated consistently strong slow wave features across nearly all metrics, including high counts, density, amplitude, and steep slopes. Like the blue cluster, the durations were relatively short, reflecting efficient oscillatory activity. This pattern matches well with prior work showing that younger and healthier populations generate more abundant and higher-amplitude slow waves with shorter durations compared to older or less healthy groups.^42,47^

Taken together, these results confirm that the identified clusters differ meaningfully in their slow wave activity during NREM sleep. Importantly, the observed patterns align closely with the expected physiological profiles of each group. This independent validation strongly supports the robustness of the four clusters, demonstrating that they capture genuine, biologically relevant differences in sleep microarchitecture rather than artifacts of the original feature space.

## DISCUSSION

We reveal distinct clusters of individuals based on sleep quality and related health metrics, offering novel insights into the relationship between sleep patterns and physical and mental health. Previous research has primarily focused on sleep disorders as categorical diagnoses, such as OSA or insomnia, often overlooking the nuanced interplay of various sleep metrics and their impact on overall health. This study’s identification of four distinct clusters—mentally healthy individuals with good deep sleep, a poor deep sleep group, a physically unhealthy group, and younger, physically healthy group—adds a critical dimension to our understanding of sleep health, suggesting that sleep quality is not merely a binary state, but a spectrum influenced by multiple interrelated factors.

The awake period immediately preceding sleep represents a transitional neural state that may provide insight into both sleep-onset dynamics and the physiological conditions that shape subsequent sleep. Spectral activity during this period, particularly within the alpha and theta bands, has been associated with changes in arousal, attention, and sleep initiation, and may relate to later sleep architecture.^50,51^ Elevated pre-sleep alpha activity, for example, has been linked to cortical hyperarousal and difficulty initiating sleep in individuals with insomnia.^52^ More broadly, waking EEG immediately before sleep may reflect cognitive and physiological processes such as attention, relaxation, and readiness for sleep, thereby providing a window into the neural state from which sleep emerges.^53^

We restricted EEG features used for clustering to pre-sleep wake to avoid circularity: sleep-stage EEG power, particularly delta power during N3, is intrinsically coupled to the macroarchitectural variables already included in the feature set. However, none of the retained pre-sleep spectral measures differed across clusters after false-discovery-rate correction. Accordingly, the present results do not support strong claims that these clusters capture distinct pre-sleep cortical states. The null finding may reflect the limited spatial sampling of a single central derivation, variable duration and timing of pre-sleep wake, the uncontrolled home environment, or the possibility that waking spectral traits contribute less to these phenotypes than demographic/clinical and sleep-stage measures. Future work should test standardized eyes-open and eyes-closed recordings, regional EEG features, spectral slope, and individualized alpha measures.

A unique strength of the SHHS dataset lies in its tracking of many subjects over an extended period of 5.2 years, allowing for a comprehensive analysis of the robustness of sleep-related clustering. Despite the observed transitions of individuals between clusters over time, the same four distinct clusters consistently emerged in both datasets, each with a similar distribution of participants. The persistence of these clusters, even as some individuals shift between them, indicates that the identified sleep profiles capture fundamental differences in sleep architecture and health status that are stable at the population level. This long-term consistency highlights the power of clustering as a tool for uncovering reliable, population-wide patterns in sleep health, offering valuable insights for future research and potential applications in personalized sleep interventions.^18,54^ Moreover, the significant transition of subjects between clusters over time underscores the dynamic nature of sleep health and the potential for change through lifestyle or therapeutic interventions.^55^

While normative studies provide valuable benchmarks for sleep metrics, there is a pressing need to consider the interplay of these metrics through cluster analysis. The four-cluster structure identified here provided a robust representation of the variation in both sleep and demographic characteristics, offering a comprehensive understanding of sleep patterns.^44,56^ Cluster analysis revealed significant differences in sleep architecture, specifically concerning N3 sleep, which is essential for cognition and memory and metabolic health.^57,58^ The clustering of subjects into groups based on N3 sleep metrics emphasizes the impact of deep sleep on physical health outcomes.^59^ The present study’s identification of a large cluster characterized by low N3 counts, high N1 counts, and comparatively poorer physical-health indicators could link poor deep sleep with adverse health outcomes such as obesity and cardiovascular disease.^60^

The artifact detection formula used here was derived empirically through systematic inspection of manually labeled artifact segments in the SHHS dataset. By comparing the spectral profiles of noise contaminated versus clean epochs, we identified consistent patterns of elevated low-frequency drift and high-frequency noise relative to mid-frequency activity. To formalize these observations, we conducted an exhaustive search to determine combinations which provided the strongest discrimination between artifact and non-artifact epochs. The resulting formula (*see Methods*) therefore operationalizes empirical observations into a reproducible automated procedure, offering a principled and interpretable approach to artifact rejection that captures both low-frequency drift and high-frequency muscle activity. To our knowledge, this represents the first application of such a frequency-band ratio approach for automated artifact detection in large-scale sleep EEG datasets and could be utilized to standardize PSG sleep-scoring and spectral analysis.

Validating the clusters using an independent slow wave analysis is crucial for ensuring the robustness and reliability of the identified subgroups, as it provides an objective means of confirming whether the clusters truly reflect underlying differences in sleep physiology. Slow waves, which are characteristic of NREM sleep stages N2 and N3, are known to be critical markers of sleep depth and restorative processes, playing a key role in synaptic homeostasis and memory consolidation.^61^ By evaluating slow wave activity, we independently assessed whether the clusters, originally derived from awake-state EEG data, correspond to meaningful variations in sleep architecture during deeper sleep stages. This validation step is particularly important as it uses a distinct set of data—occurring during NREM periods—entirely separate from the awake EEG data used in the cluster analysis. Such independent validation is supported in the literature, as previous studies have highlighted the importance of using multiple, independent measures to validate sleep-related subgroups and ensure that identified clusters are not artifacts of any single metric or time period.^8^

Our findings highlight that older individuals tend to cluster into groups with reduced deep sleep (N3) and slow-wave activity, which aligns with previous research indicating that sleep fragmentation and the decline of slow-wave sleep are common in aging.^8,30^ This decline in slow-wave activity has critical implications, as it is associated with reduced restorative processes, such as memory consolidation and neuroplasticity, both of which are essential for maintaining cognitive health in older adults.^61,62^ In contrast, the robust slow wave activity observed in Clusters 1 and 4 may indicate resilience factors or cognitive protective mechanisms that might correlate with healthier sleep architecture.^29,63^

The identification of distinct clusters and slow wave characteristics raises critical implications for personalized sleep medicine using non-pharmacological approaches, particularly brain-computer interfaces (BCIs) aimed at enhancing sleep quality by recognizing and responding to individual variations in sleep micro- and macroarchitectures. By recognizing an individual’s specific sleep profile cluster, BCIs could dynamically adjust intervention thresholds and protocols to align with personal baseline patterns, enhancing effectiveness. These systems could also learn from long-term EEG data, adapting intervention strategies over time; for instance, if a decline in slow-wave activity is consistently detected, the BCI could alter or increase intervention intensity. This adaptive, personalized approach has the potential to not only improve immediate sleep quality but also support long-term health by promoting restorative sleep phases, ultimately creating BCIs that move beyond tracking-only to active, individualized sleep management systems.

Wearable BCI devices equipped with real-time EEG could monitor sleep architecture and slow-wave activity relative to individualized baseline profiles. During N2 and N3 sleep, closed-loop algorithms could detect ongoing slow oscillations and deliver precisely timed auditory stimulation to enhance slow-wave activity, synchronization, and sleep-dependent memory processes.^64–66^ Because slow-wave sleep is associated with glymphatic transport and metabolic waste clearance, such systems could also test whether improved slow-wave physiology produces corresponding changes in clearance-related biomarkers.^62^ Pre-sleep EEG could further identify hyperarousal or elevated high-frequency activity associated with impaired sleep initiation and trigger individualized relaxation or sensory-conditioning protocols.^67,68^ Finally, the system could track REM continuity and sleep-cycle progression, adapting or suspending stimulation when necessary to avoid disrupting REM-dependent emotional and cognitive functions.^69^

In conclusion, this study’s identification of separate clusters of individuals based on sleep quality metrics not only adds depth to the current understanding of sleep health but also paves the way for more nuanced and personalized approaches to clinical sleep medicine. By highlighting the intricate relationships among sleep architecture, health perceptions, and clinical characteristics, these findings encourage further exploration into the multifaceted nature of sleep and its critical role in overall well-being.

### Study Limitations

Several limitations should be considered. Sleep stages were manually scored and therefore remain subject to inter-scorer variability, although the SHHS used centralized scoring and quality-control procedures. The analysis used a single central EEG derivation for spectral and slow-wave measures, limiting assessment of regional topography. Pre-sleep wake was not collected as a standardized resting-state protocol and varied in duration and behavioral context; this likely reduced sensitivity to stable waking EEG differences. Feature selection and cluster discovery were performed in SHHS-1 and evaluated in the same longitudinal cohort at SHHS-2 rather than in a fully independent external cohort. In addition, exclusion of participants with missing, zero, or extreme values produced a complete-case sample that may not represent the full SHHS population. Cluster labels are descriptive summaries of relative feature patterns and should not be interpreted as diagnoses or discrete biological subtypes.

The analysis of cluster transfers over time does not establish causal relationships between sleep metrics and health outcomes. Continuous, longitudinal controlled studies would be necessary to determine the directionality of these associations and to explore how changes in sleep patterns might impact health over time. Understanding these causal relationships could further inform targeted interventions and enhance our understanding of the mechanisms underlying the observed sleep clusters.

## Data Availability

All data produced in the present study are available upon reasonable request to the authors.

https://sleepdata.org/datasets/shhs

## DISCLOSURE STATEMENT

The authors declare the following financial interests/personal relationships which may be considered as potential competing interests: A.P., K.L.M., and C.H.G. have received salaries from, and have equity/stock options in, Attune Neurosciences, Inc.; A.P. and C.H.G. (Founder CEO) are employed by Thalamix Labs. The opinions or assertions contained herein are the views of the authors and are not to be construed as the views of the U.S. Department of Defense, Walter Reed National Military Medical Center (WRNMMC), or the Uniformed Services University (USU).

## FUNDING

This work was supported by the United States Department of Defense under Award/Contract No. W81XWH-22-C-0018.

## Notes

### Author Declarations

The Sleep Heart Health Study protocol was approved by the institutional review board of each participating institution, and all participants provided written informed consent. The present study was a secondary analysis of existing Sleep Heart Health Study data obtained through the National Sleep Research Resource under an approved Data Access and Use Agreement. No additional participants were recruited and no new data were collected for this analysis.

