## Supplemental Tables and Figures for "Longitudinal Sleep-Health Phenotypes Identified by Hierarchical Clustering in the Sleep Heart Health Study"

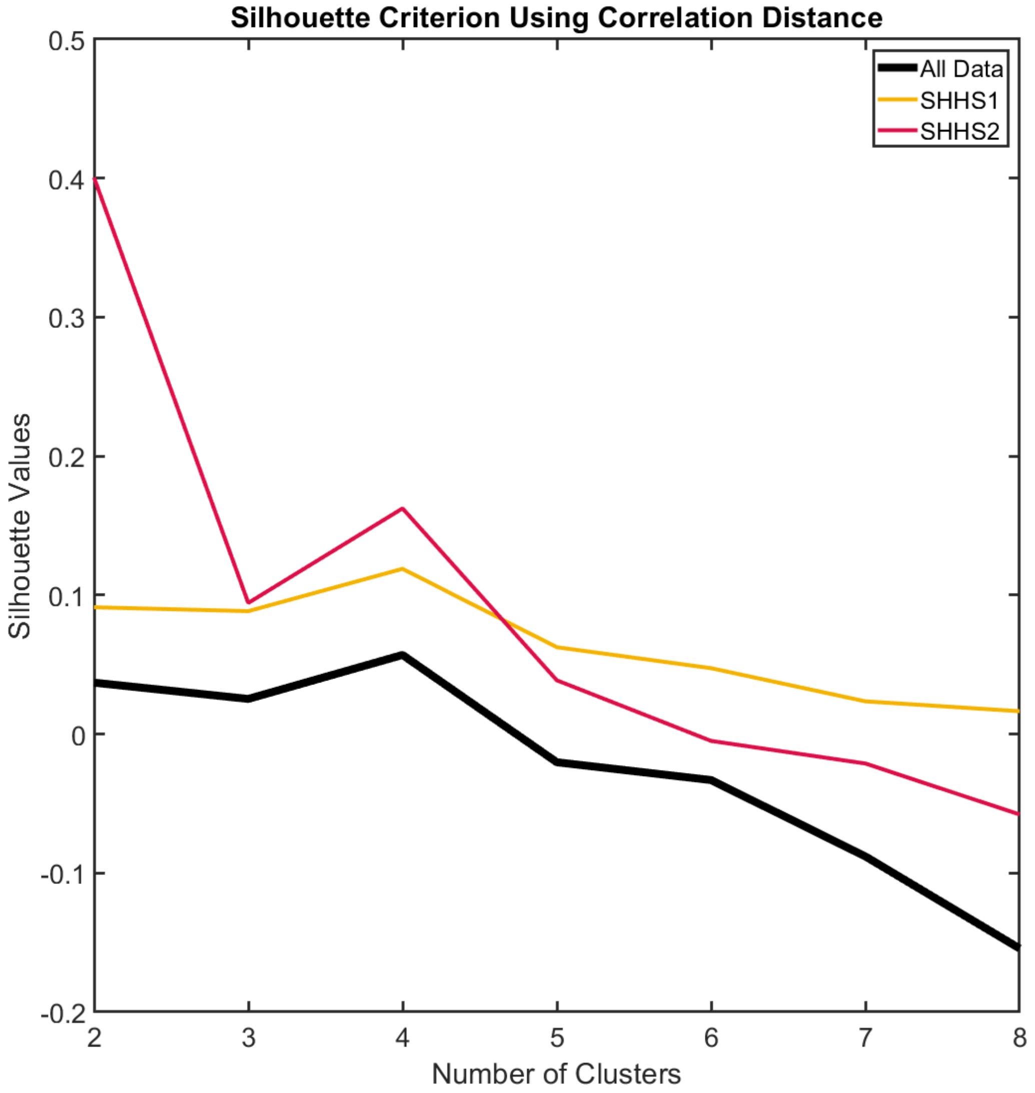

**Supplemental Figure 1. Silhouette Criterion for Cluster Optimization in SHHS-1, SHHS-2, and Combined Datasets.** The silhouette criterion was used to determine the optimal number of clusters for SHHS-1 (orange), SHHS-2 (red), and the combined SHHS-1+SHHS-2 dataset (black). Silhouette scores for different numbers of clusters (ranging from 2 to 8) are plotted on the y-axis, with the number of clusters on the x-axis, providing insight into the clustering stability across different configurations. For SHHS-1 and the combined dataset, four clusters were identified as optimal, as indicated by the highest average silhouette score. For SHHS-2, two clusters showed the highest silhouette score, though four clusters ranked as the second-best solution. This suggests that, while the SHHS-2 data may have a slightly different structure, the four-cluster solution remains robust across datasets.

**A.**

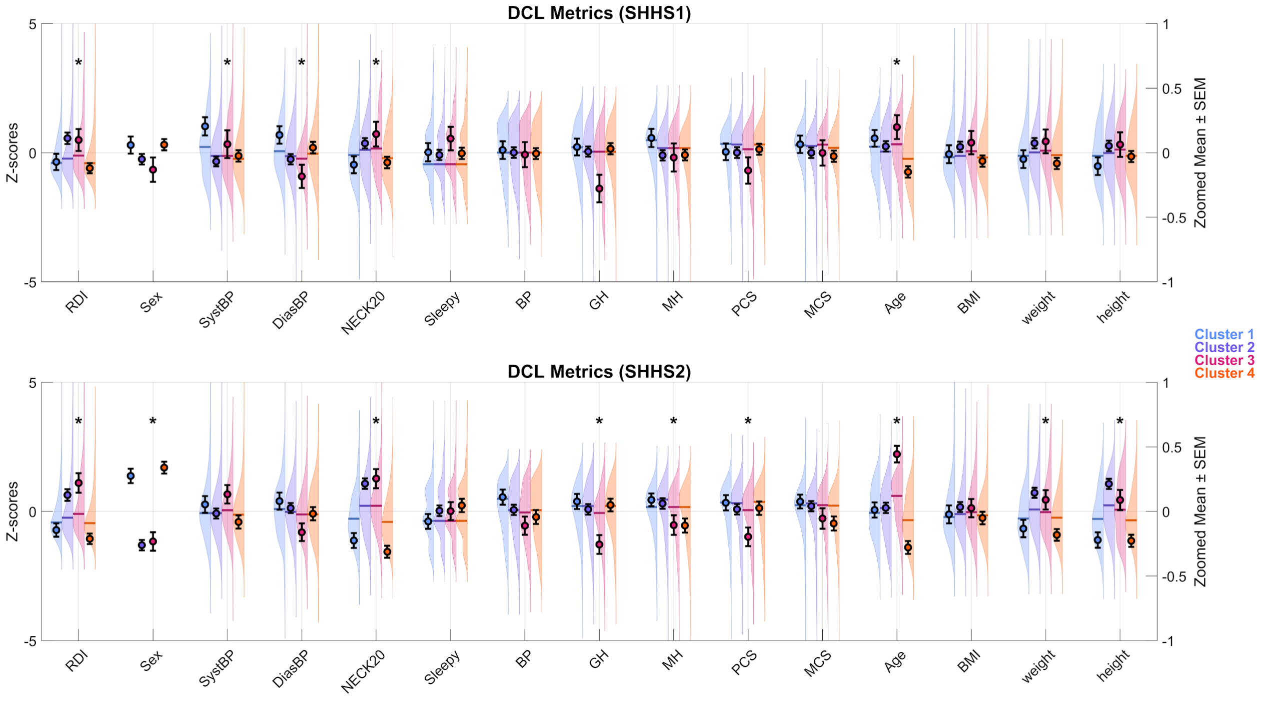
DCL Metrics (SHHS1)
Z-scores
Zoomed Mean ± SEM
5
0
-5
1
0.5
0
-0.5
-1
RDI
Sex
SystBP
DiasBP
NECK20
Sleepy
BP
GH
MH
PCS
MCS
Age
BMI
weight
height
Cluster 1
Cluster 2
Cluster 3
Cluster 4
DCL Metrics (SHHS2)
Z-scores
Zoomed Mean ± SEM
RDI
Sex
SystBP
DiasBP
NECK20
Sleepy
BP
GH
MH
PCS
MCS
Age
BMI
weight
height

**B.**

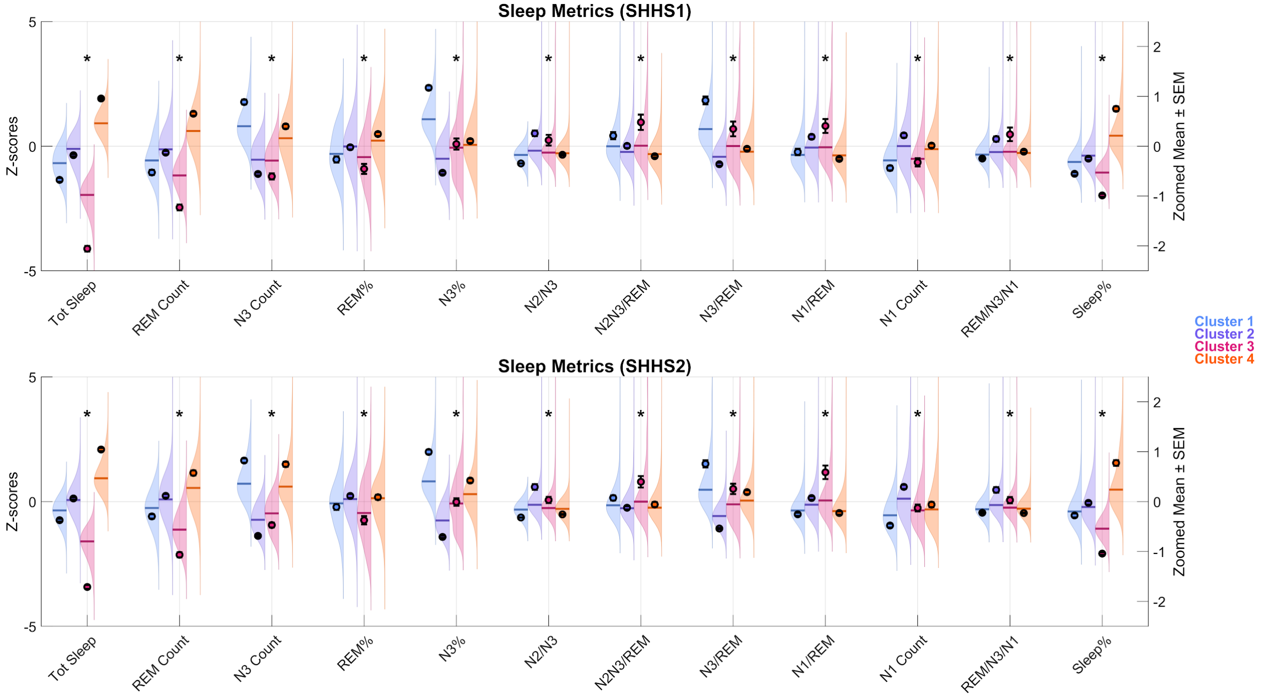
Sleep Metrics (SHHS1)
Z-scores
Zoomed Mean ± SEM
5
0
-5
2
1
0
-1
-2
Tot Sleep
REM Count
N3 Count
REM%
N3%
N2/N3
N2N3/REM
N3/REM
N1/REM
N1 Count
REM/N3/N1
Sleep%
Cluster 1
Cluster 2
Cluster 3
Cluster 4
Sleep Metrics (SHHS2)
Z-scores
Zoomed Mean ± SEM
Tot Sleep
REM Count
N3 Count
REM%
N3%
N2/N3
N2N3/REM
N3/REM
N1/REM
N1 Count
REM/N3/N1
Sleep%

C.

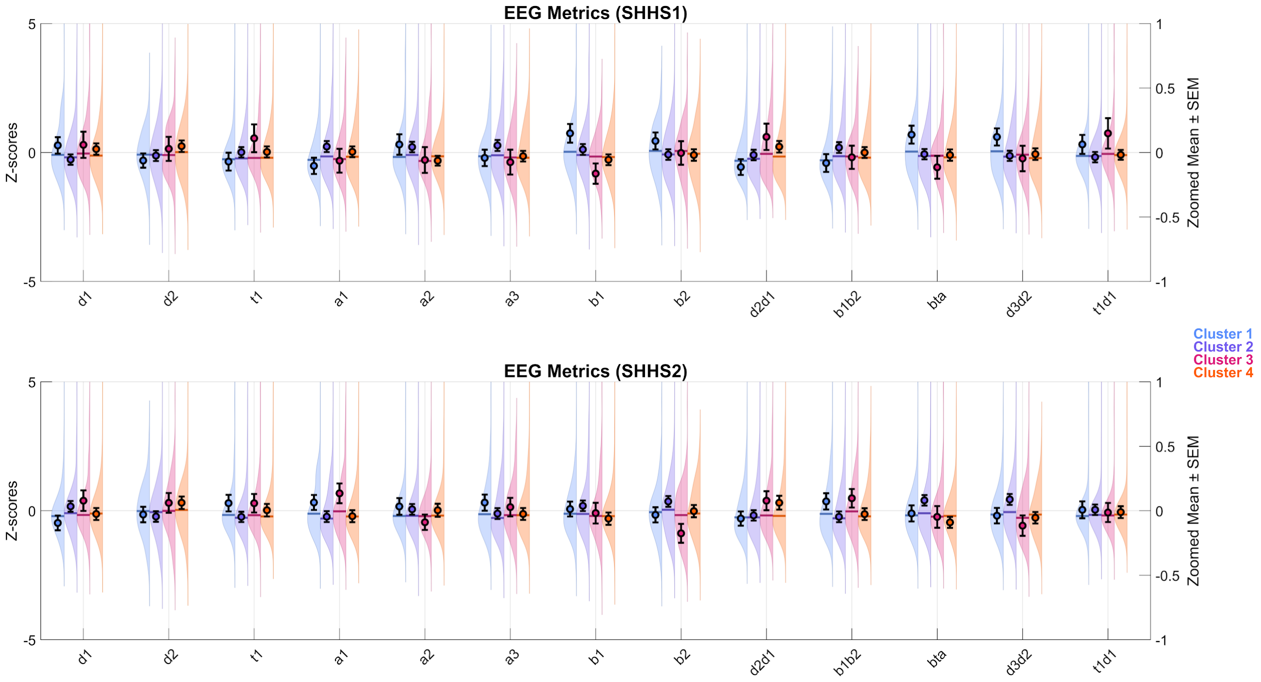

**Supplemental Figure 2. Cluster differences across health, sleep, and EEG domains.** (A) DCL metrics include demographic, clinical, and lifestyle measures. (B) Sleep metrics reflect polysomnography-derived architecture and continuity features. (C) EEG metrics capture spectral power across canonical frequency bands. Violin plots show the distribution of values for each cluster in SHHS1 (top) and SHHS2 (bottom), expressed as z-scores. Black points and error bars represent cluster means ± SEM on a zoomed scale. Clusters are color-coded (Cluster 1 = blue, Cluster 2 = purple, Cluster 3 = pink, Cluster 4 = orange). Asterisks (*) denote significant cluster differences after one-way ANOVA with false discovery rate (FDR) correction ($q < 0.05$).

**Supplemental Table 1. DCL, PSG, and EEG metrics averaged across clusters in SHHS-1.** Summary of the average values (with interquartile ranges or counts where applicable) of 40 key demographic, clinical, PSG, and EEG measures for each of the four clusters. Bolded features indicate statistically significant differences across clusters based on ANOVA, with p-values corrected for multiple comparisons using FDR.

| | Feature | Cluster 1 (n = 227) | Cluster 2 (n = 606) | Cluster 3 (n = 114) | Cluster 4 (n = 521) | units |
|---|---|---|---|---|---|---|
| **1** | **RDIsn** | **7.12 [0.8;13.4]** | **9.22 [2;16.4]** | **10.65 [3.2;18.1]** | **7 [1.3;12.7]** | events/hr |
| 2 | sex | 133 (59) | 322 (53) | 56 (49) | 306 (59) | female count (%) |
| **3** | **SystBP** | **128 [114.5;141.5]** | **122[112;132]** | **122 [109;135]** | **120 [108.5;131.5]** | mmHg |
| **4** | **DiasBP** | **73 [66;80]** | **71 [64;78]** | **70 [65.5;74.5]** | **72 [65.5;78.5]** | mmHg |
| **5** | **NECK** | **37 [34.3;39.8]** | **37.8 [34.9;40.8]** | **38 [35.4;40.6]** | **36.5 [33.5;39.5]** | cm |
| 6 | Sleepy | 2.41 (0.97) | 2.39 (0.87) | 2.5 (0.87) | 2.4 (0.91) | scale (0-24) |
| 7 | bp | 74 [54.5;93.5] | 74 [54.5;93.5] | 74 [54.5;93.5] | 74 [62.5;85.5] | scale (10-100) |
| **8** | **gh** | **80 [68.9;91.1]** | **77 [65.5;88.5]** | **77 [63.5;90.5]** | **77 [65.5;88.5]** | scale (10-100) |
| 9 | mh | 88 [80;96] | 84 [76;92] | 84 [74;94] | 84 [76;92] | scale (10-100) |
| 10 | pcs | 51.92 [46.1;57.7] | 51.7 [46.5;56.9] | 50.07 [43.5;56.6] | 51.71 [46.6;56.9] | scale (10-100) |
| 11 | mcs | 56.45 [53;59.9] | 56.26 [52.7;59.9] | 56.64 [52.7;60.5] | 55.73 [52;59.4] | scale (10-100) |
| **12** | **age** | **65 [56.6;73.4]** | **63 [55;71]** | **66 [58;74]** | **60 [52.4;67.6]** | years |
| 13 | bmi | 27.16 [24.1;30.2] | 27.41 [24.5;30.3] | 28.26 [25.5;31] | 27.12 [24.1;30.1] | kg/m² |
| 14 | weight | 75.5 [65.5;85.5] | 77.5 [67.6;87.5] | 78.5 [70.6;86.5] | 75.9 [65.4;86.4] | kg |
| 15 | height | 166 [158.4;173.7] | 167 [159.9;174.2] | 168.25 [161.8;174.8] | 166 [159;173] | cm |
| **16** | **Tot Sleep** | **653 [603.6;702.4]** | **720 [674;766]** | **504 [459;549]** | **840 [805.8;874.3]** | half minutes |
| **17** | **REM Count** | **122 [97.3;146.8]** | **145 [115.5;174.5]** | **91 [69.5;112.5]** | **183 [152;214]** | half minutes |
| **18** | **N3 Count** | **186 [152.5;219.5]** | **88.5 [49.5;127.5]** | **86 [48.5;123.5]** | **151 [103.9;198.1]** | half minutes |
| **19** | **REM%** | **0.19 [0.1;0.2]** | **0.2 [0.2;0.2]** | **0.18 [0.1;0.2]** | **0.22 [0.2;0.3]** | percent |
| **20** | **N3%** | **0.28 [0.2;0.3]** | **0.13 [0.1;0.2]** | **0.17 [0.1;0.3]** | **0.18 [0.1;0.2]** | percent |
| **21** | **N2/N3** | **1.66 [1.2;2.1]** | **4.82 [1.4;8.3]** | **3.42 [0.6;6.2]** | **2.93 [1.6;4.3]** | ratio |
| **22** | **N2N3/REM** | **4.14 [3.1;5.2]** | **3.6 [2.7;4.5]** | **4.2 [2.9;5.5]** | **3.39 [2.6;4.2]** | ratio |
| **23** | **N3/REM** | **1.53 [1.1;1.9]** | **0.63 [0.4;0.9]** | **0.98 [0.5;1.5]** | **0.8 [0.5;1.1]** | ratio |
| **24** | **N1/REM** | **0.19 [0.1;0.3]** | **0.27 [0.1;0.4]** | **0.27 [0.1;0.4]** | **0.18 [0.1;0.3]** | ratio |
| **25** | **N1 Count** | **23 [13;33]** | **37 [21.5;52.5]** | **24.5 [12.5;36.5]** | **34 [20.5;47.5]** | half minutes |
| **26** | **REM/N3/N1** | **0.03 [0;0]** | **0.05 [0;0.1]** | **0.05 [0;0.1]** | **0.04 [0;0.1]** | ratio |
| **27** | **Sleep/wake** | **1.94 [1.5;2.4]** | **2.52 [1.9;3.1]** | **0.96 [0.8;1.1]** | **4.37 [3.2;5.5]** | percentage |

| | | | | | | |
|---|---|---|---|---|---|---|
| 28 | d1 | 3.9 [3.1;4.7] | 3.76 [3;4.6] | 3.96 [3.1;4.8] | 3.85 [3.1;4.6] | μV/Hz |
| 29 | d2 | 2.97 [2.6;3.4] | 2.97 [2.5;3.5] | 2.96 [2.4;3.5] | 3.04 [2.5;3.6] | μV/Hz |
| 30 | t1 | 1.04 [0.9;1.2] | 1.05 [0.9;1.2] | 1.06 [0.9;1.2] | 1.06 [0.9;1.2] | μV/Hz |
| 31 | a1 | 1.08 [0.9;1.3] | 1.13 [0.9;1.3] | 1.09 [0.9;1.3] | 1.13 [0.9;1.3] | μV/Hz |
| 32 | a2 | 0.91 [0.8;1] | 0.92 [0.8;1] | 0.88 [0.8;1] | 0.91 [0.8;1] | μV/Hz |
| 33 | a3 | 1.01 [0.9;1.1] | 1.02 [0.9;1.2] | 1 [0.9;1.1] | 1 [0.9;1.1] | μV/Hz |
| **34** | **b1** | **0.81 [0.7;0.9]** | **0.8 [0.7;0.9]** | **0.79 [0.7;0.9]** | **0.79[0.7;0.9]** | μV/Hz |
| 35 | b2 | 0.59 [0.5;0.7] | 0.57 [0.5;0.7] | 0.59 [0.5;0.7] | 0.57 [0.5;0.7] | μV/Hz |
| 36 | d2d1 | 0.07 [0;0.1] | 0.07 [0;0.1] | 0.08 [0;0.1] | 0.08 [0;0.1] | ratio |
| 37 | b1b2 | 1.54[ 1.2;1.9] | 1.62 [1.3;2] | 1.62 [1.3;1.9] | 1.59 [1.3;1.9] | ratio |
| 38 | bta | 0.71 [0.6;0.9] | 0.67 [0.5;0.8] | 0.67 [0.5;0.8] | 0.66 [0.5;0.8] | ratio |
| 39 | d3d2 | 0.63 [0.5;0.8] | 0.58 [0.4;0.7] | 0.6 [0.5;0.7] | 0.57 [0.4;0.7] | ratio |
| 40 | t1d1 | 1.71 [1.5;1.9] | 1.71 [1.5;1.9] | 1.75 [1.5;2] | 1.72 [1.5;1.9] | ratio |

**Supplemental Table 2. DCL, PSG, and EEG metrics averaged across clusters in SHHS-2.** Summary of the average values (with interquartile ranges or counts where applicable) of 40 key demographic, clinical, PSG, and EEG measures for each of the four clusters. Bolded features indicate statistically significant differences across clusters based on ANOVA, with p-values corrected for multiple comparisons using the FDR.

| | Feature | Cluster 1 (n = 277) | Cluster 2 (n = 614) | Cluster 3 (n = 195) | Cluster 4 (n = 382) | units |
|---|---|---|---|---|---|---|
| **1** | **RDIsn** | **9.01 [2;16]** | **11.67 [2.8;20.6]** | **13.82 [3.2;24.4]** | **8.6 [2.5;14.7]** | events/hr |
| **2** | **sex** | **192 (0.69)** | **262 (0.43)** | **86 (0.44)** | **277 (0.73)** | female count (mean) |
| 3 | SystBP | 126 [115;137] | 126 [115.5;136.5] | 128 [118;138] | 125 [114.5;135.5] | mmHg |
| 4 | DiasBP | 71 [64.4;77.6] | 70 [63.5;76.5] | 69 [63;75] | 69 [62;76] | mmHg |
| **5** | **NECK20** | **36 [33.3;38.8]** | **38 [35;41]** | **38 [35.6;40.4]** | **35.5 [33;38]** | cm |
| 6 | Sleepy | 2.25 (0.83) | 2.32 (0.89) | 2.32 (0.84) | 2.36 (0.87) | scale (0-24) |
| 7 | bp | 84 [65;103] | 74 [62.5;85.5] | 72 [60.1;83.9] | 74 [49.5;98.5] | scale (10-100) |
| **8** | **gh** | **77 [64.5;89.5]** | **77 [64.5;89.5]** | **72 [59.8;84.3]** | **77 [66;88]** | scale (10-100) |
| **9** | **mh** | **84 [77.5;90.5]** | **88 [80;96]** | **84 [74;94]** | **84 [74;94]** | scale (10-100) |
| 10 | pcs | 50.43 [44;56.9] | 50.14 [44.1;56.2] | 47.39 [40.7;54] | 50.67 [43.7;57.6] | scale (10-100) |
| 11 | mcs | 56.77 [53.3;60.2] | 57.3 [54.1;60.5] | 56.78 [52.5;61.1] | 56.67 [52;61.3] | scale (10-100) |
| **12** | **age** | **67 [59;75]** | **69 [61;77]** | **74 [66.6;81.4]** | **64 [56;72]** | years |
| 13 | bmi | 27.05 [23.6;30.5] | 27.6 [24.8;30.4] | 27.97 [24.6;31.3] | 27.37 [24.5;30.2] | kg/m² |
| **14** | **weight** | **74.4 [62.6;86.2]** | **79.95 [69.5;90.4]** | **78.3 [67.1;89.5]** | **74.9 [65.3;84.5]** | kg |
| **15** | **height** | **163 [156.1;169.9]** | **168 [161;175]** | **166.4 [157.8;175.1]** | **162.5 [156.6;168.5]** | cm |
| **16** | **Tot Sleep** | **706 [666.4;745.6]** | **760 [714.5;805.5]** | **545 [498.5;591.5]** | **873 [825;921]** | half mintues |
| **17** | **REM Count** | **146 [119.1;172.9]** | **165.5 [133;198]** | **97 [69.1;124.9]** | **191.5 [156.5;226.5]** | half mintues |
| **18** | **N3 Count** | **175 [138.5;211.5]** | **63 [31;95]** | **83 [42;124]** | **166 [127;205]** | half mintues |
| **19** | **REM%** | **0.21 [0.2;0.2]** | **0.22 [0.2;0.3]** | **0.18 [0.1;0.2]** | **0.22 [0.2;0.3]** | percent |
| **20** | **N3%** | **0.24 [0.2;0.3]** | **0.08 [0;0.1]** | **0.15 [0.1;0.2]** | **0.19 [0.1;0.2]** | percent |
| **21** | **N2/N3** | **2.04 [1.5;2.6]** | **7.49 [1.9;13]** | **3.76 [0.6;6.9]** | **2.87 [1.8;3.9]** | ratio |
| **22** | **N2N3/REM** | **3.62 [2.7;4.6]** | **3.3 [2.6;4]** | **4.02 [2.6;5.4]** | **3.36 [2.6;4.1]** | ratio |
| **23** | **N3/REM** | **1.24 [0.8;1.7]** | **0.4 [0.2;0.6]** | **0.78 [0.3;1.3]** | **0.9 [0.6;1.2]** | ratio |
| **24** | **N1/REM** | **0.19 [0.1;0.3]** | **0.27 [0.2;0.4]** | **0.33 [0.2;0.5]** | **0.18 [0.1;0.3]** | ratio |
| **25** | **N1 Count** | **27 [16;38]** | **44 [28.5;59.5]** | **32 [18.5;45.5]** | **33 [17.5;48.5]** | half minutes |
| **26** | **REM/N3/N1** | **0.03 [0;0]** | **0.07 [0;0.1]** | **0.04 [0;0.1]** | **0.04 [0;0.1]** | ratio |

| | | | | | | |
|---|---|---|---|---|---|---|
| **27** | **Sleep/wake** | **1.56 [1.2;1.9]** | **1.73 [1.2;2.2]** | **0.88 [0.7;1.1]** | **2.42 [1.8;3.1]** | percentage |
| 28 | d1 | 3.37 [2.8;4] | 3.52 [2.9;4.2] | 3.43 [2.8;4.1] | 3.5 [2.9;4.1] | µV/Hz |
| 29 | d2 | 3.08 [2.6;3.6] | 3.04 [2.6;3.5] | 3.08 [2.6;3.6] | 3.11 [2.6;3.6] | µV/Hz |
| 30 | t1 | 1.1 [1;1.2] | 1.07 [1;1.2] | 1.1 [1;1.2] | 1.08 [1;1.2] | µV/Hz |
| 31 | a1 | 1.02 [0.9;1.2] | 0.97 [0.8;1.1] | 1.05 [0.9;1.2] | 0.99 [0.9;1.1] | µV/Hz |
| 32 | a2 | 0.88 [0.8;1] | 0.88 [0.8;1] | 0.87 [0.8;1] | 0.88 [0.8;1] | µV/Hz |
| 33 | a3 | 0.95 [0.8;1.1] | 0.93 [0.8;1.1] | 0.95 [0.8;1.1] | 0.95 [0.8;1] | µV/Hz |
| 34 | b1 | 0.84[0.7;0.9] | 0.84[0.7;0.9] | 0.83[0.7;0.9] | 0.83 [0.7;0.9] | µV/Hz |
| **35** | **b2** | **0.54[0.5;0.6]** | **0.55[0.5;0.6]** | **0.53[0.4;0.6]** | **0.53 [0.5;0.6]** | µV/Hz |
| **36** | d2/d1 | 0.08 [0.1;0.1] | 0.08 [0.1;0.1] | 0.09 [0.1;0.1] | 0.09 [0.1;0.1] | ratio |
| 37 | b1/b2 | 1.55 [1.2;1.8] | 1.47 [1.2;1.8] | 1.59 [1.3;1.9] | 1.5 [1.3;1.7] | ratio |
| 38 | bta | 0.65 [0.5;0.8] | 0.67 [0.5;0.8] | 0.64 [0.5;0.8] | 0.64 [0.5;0.8] | ratio |
| 39 | d3/d2 | 0.53 [0.4;0.6] | 0.55 [0.4;0.7] | 0.51 [0.4;0.6] | 0.53 [0.4;0.6] | ratio |
| 40 | t1/d1 | 1.78 [1.6;2] | 1.81 [1.6;2] | 1.79 [1.6;2] | 1.82 [1.6;2] | ratio |

**Supplemental Table 3. ANOVA results of slow wave metrics across clusters from SHHS-1 and SHHS-2.** One-way analyses of variance were conducted separately within each dataset to evaluate differences among the four clusters for eight slow-wave metrics: count, density, absolute negative amplitude, total amplitude, total duration, negative duration, downslope, and upslope. Raw *p*-values were corrected across all slow-wave comparisons using the false discovery rate procedure. All eight metrics differed significantly across clusters in both SHHS-1 and SHHS-2 after FDR correction ($q < 0.05$).

| Dataset | Metric | F_stat | p_raw | p_fdr_global | FDR < 0.05 |
|---|---|---|---|---|---|
| **SHHS1** | Count | 48.03 | 1.34E-29 | 7.17E-29 | TRUE |
| | Density | 42.96 | 1.33E-26 | 5.34E-26 | TRUE |
| | Negative Amplitude (abs) | 14.56 | 2.40E-09 | 3.84E-09 | TRUE |
| | Total Amplitude | 19.68 | 1.67E-12 | 5.34E-12 | TRUE |
| | Duration | 5.16 | 0.0015124 | 0.0015124 | TRUE |
| | Negative Duration | 5.59 | 0.000827981 | 0.000883179 | TRUE |
| | DownSlope | 11.49 | 1.92E-07 | 2.56E-07 | TRUE |
| | UpSlope | 14.41 | 2.99E-09 | 4.35E-09 | TRUE |
| **SHHS2** | Count | 196.57 | 4.11E-107 | 6.58E-106 | TRUE |
| | Density | 130.66 | 5.34E-75 | 4.27E-74 | TRUE |
| | Negative Amplitude (abs) | 16.06 | 2.84E-10 | 6.49E-10 | TRUE |
| | Total Amplitude | 18.23 | 1.29E-11 | 3.44E-11 | TRUE |
| | Duration | 11.43 | 2.08E-07 | 2.56E-07 | TRUE |
| | Negative Duration | 10.73 | 5.67E-07 | 6.48E-07 | TRUE |
| | DownSlope | 14.61 | 2.23E-09 | 3.84E-09 | TRUE |
| | UpSlope | 15.18 | 9.85E-10 | 1.97E-09 | TRUE |
